# Salbutamol, a short acting beta-2 agonist, reduces risk and improves prognosis of prostate cancer

**DOI:** 10.1101/2024.02.16.24302956

**Authors:** Steven Lehrer, Peter H. Rheinstein

## Abstract

**Background:** Beta-blockers, a class of drugs commonly used to manage blood pressure, have been the subject of research regarding their relationship to prostate cancer risk, prognosis, and treatment. Beta blockers reduce risk and improve prognosis of prostate cancer. Perioperative use of a non-selective beta blocker improves outcome after radical prostatectomy. But a related class of drugs, beta 2 adrenergic agonists, has received little attention in prostate cancer.

**Methods:** We studied the relationship of the beta 2 adrenergic agonist salbutamol to prostate cancer risk and survival. We analyzed FDA MedWatch data to determine whether salbutamol could influence the risk of prostate cancer. We used UK Biobank (UKBB) data to assess the effect of salbutamol on prostate cancer (PC) survival.

**Results:** Salbutamol significantly reduces prostate cancer risk, Proportional Reporting Ratio (PRR) and 95% confidence interval (lower bound; upper bound): 0.131 (0.11; 0.155) and improves prognosis. Mean survival was 7.35 years for subjects not taking salbutamol, 10.5 years for subjects taking salbutamol (p = 0.041, log rank test. To adjust for the effect of age we performed proportional hazards regression, survival time dependent variable, age and salbutamol use independent variables. Salbutamol use was significantly related to survival time (p = 0.016) and independent of the significant effect of age (p < 0.001).

**Conclusion:** Salbutamol and other beta-adrenergic agonists could represent a new class of drugs for treatment of prostate cancer.

---

Prostate cancer is the second most common cancer in men worldwide and a leading cause of cancer-related deaths. Despite advancements in screening and treatment, a need remains for novel therapeutic strategies to improve outcomes for patients with prostate cancer. Beta-blockers, a class of medications that antagonize beta-adrenergic receptors, have attracted attention due to their potential role in cancer prevention and treatment beyond their cardiovascular effects.

Beta-blockers, commonly used to manage blood pressure, have been the subject of research in prostate cancer risk and treatment. Beta blockers reduce prostate cancer risk and improve outcome (1,2). Perioperative use of a non-selective beta blocker improves prognosis after radical prostatectomy (3). The beta blocker carvedilol is currently being tested in this regard.

Beta-blockers, besides regulating processes such as blood pressure, heart rate, and airway strength or reactivity, interfere with processes that trigger tumorigenesis, angiogenesis, and metastasis.

The potential anticancer effects of beta-blockers are mediated through several mechanisms, including inhibition of adrenergic signaling, modulation of the immune response, and interference with angiogenesis and metastasis. Prostate cancer cells express beta-adrenergic receptors, and activation of these receptors has been linked to tumor growth, invasion, and resistance to therapy. Beta-blockers counteract these effects by blocking beta-adrenergic signaling pathways, thereby inhibiting cancer cell proliferation and promoting apoptosis (4).

A related class of drugs, beta 2 adrenergic agonists, has received little attention in prostate cancer. Beta agonists are potent bronchodilators that help manage airway narrowing in asthma and chronic obstructive pulmonary disease. Their selective action on beta-2 receptors makes them valuable tools in respiratory care.

In this article we studied the relationship of the beta 2 adrenergic agonist salbutamol to prostate cancer risk and survival. We analyzed FDA MedWatch data to determine whether salbutamol could influence the risk of prostate cancer. We used UK Biobank (UKBB) data to assess the effect of salbutamol on prostate cancer (PC) survival.

## Methods

MedWatch is the Food and Drug Administration (FDA) Safety Information and Adverse Event Reporting Program. MedWatch was organized in 1993 to collect data regarding adverse events in healthcare. An adverse event is any undesirable experience associated with the use of a medical product. The MedWatch system collects reports of adverse reactions and quality problems, primarily due to drugs and medical devices, but also for other FDA-regulated products (e.g., dietary supplements, cosmetics, medical foods, and infant formulas) (5).

Machine-readable data from MedWatch, including adverse drug reaction reports from manufacturers, are part of a public database. We used the publicly available online tool OpenVigil (6) to query the database at https://openvigil.sourceforge.net/. Results reported are the rates of the adverse event among users of a particular medication versus the rate of the adverse event among users of all other medications, as well as measures of disproportionality: observed-expected ratios like Relative Reporting Ratio, Proportional Reporting Ratio, and Reporting Odds Ratio. The Relative reporting ratio (RRR) is the ratio of how many adverse drug reactions (ADRs) under exposure were observed over the number of expected events under the assumption that ADR and drug exposure were independent. The proportional reporting ratio (PRR) is the proportion of spontaneous reports for a given drug that are linked to a specific adverse outcome, divided by the corresponding proportion for all or several other drugs. Reporting Odds Ratio (ROR) represents the odds of a certain event occurring with a medicinal product, compared to the odds of the same event occurring with all other medicinal products in the database (7). A signal is considered when the upper limit of the 95% confidence interval (CI) of the PRR, RRR, or ROR is less than one or the lower limit of the 95% confidence interval is greater than one.

The UK Biobank is a large prospective observational study of men and women with no link to MedWatch. Participants were recruited from across 22 centers located throughout England, Wales, and Scotland between 2006 and 2010 and continue to be longitudinally followed for capture of subsequent health events (8). This methodology is like that of the ongoing Framingham Heart Study (9), with the exception that the UKB program collects postmortem samples, which Framingham did not.

UK Biobank: has approval from the Northwest Multi-center Research Ethics Committee (MREC) to obtain and disseminate data and samples from the participants, and these ethical regulations cover the work in this study. Written informed consent was obtained from all participants. Details can be found at www.ukbiobank.ac.uk/ethics.

Our UK Biobank application was approved as UKB project 57245 (S.L., P.H.R.). Our analysis included all subjects with PC that occurred either before or after participant enrollment and was recorded in the UK Biobank database using self-reported data and the International Classification of Diseases (ICD10, ICD9). UK Biobank does not have date of last followup visit, so we had no censored data available, only survival, date of diagnosis to date of death. Subjects with BMI ≥ 27 were selected because treatment with drugs such as enzalutamide is effective only for patients with higher BMIs and is not effective for patients with low BMI (10).

Statistical analysis was done with SPSS v 26, (IBM, New York).

## Results

Salbutamol use is associated with a significantly reduced risk of prostate cancer (PRR 0.131). Table 1 shows MedWatch data to evaluate the relationship of salbutamol use to prostate cancer in 11,737,133 subjects. The greater the chi-squared value, the greater the differences. Chi square values greater than 3.84 are considered statistically significant. Measurements of disproportionality (observed-expected ratios RRR, PRR, ROR): Relative Reporting Ratio (RRR) and 95% confidence interval (lower bound; upper bound): 0.131 (0.112; 0.157); Proportional Reporting Ratio (PRR) and 95% confidence interval (lower bound; upper bound): 0.131 (0.11; 0.155); Reporting Odds Ratio (ROR) and 95% confidence interval (lower bound; upper bound): 0.131 (0.11; 0.154).

**Table 1.**
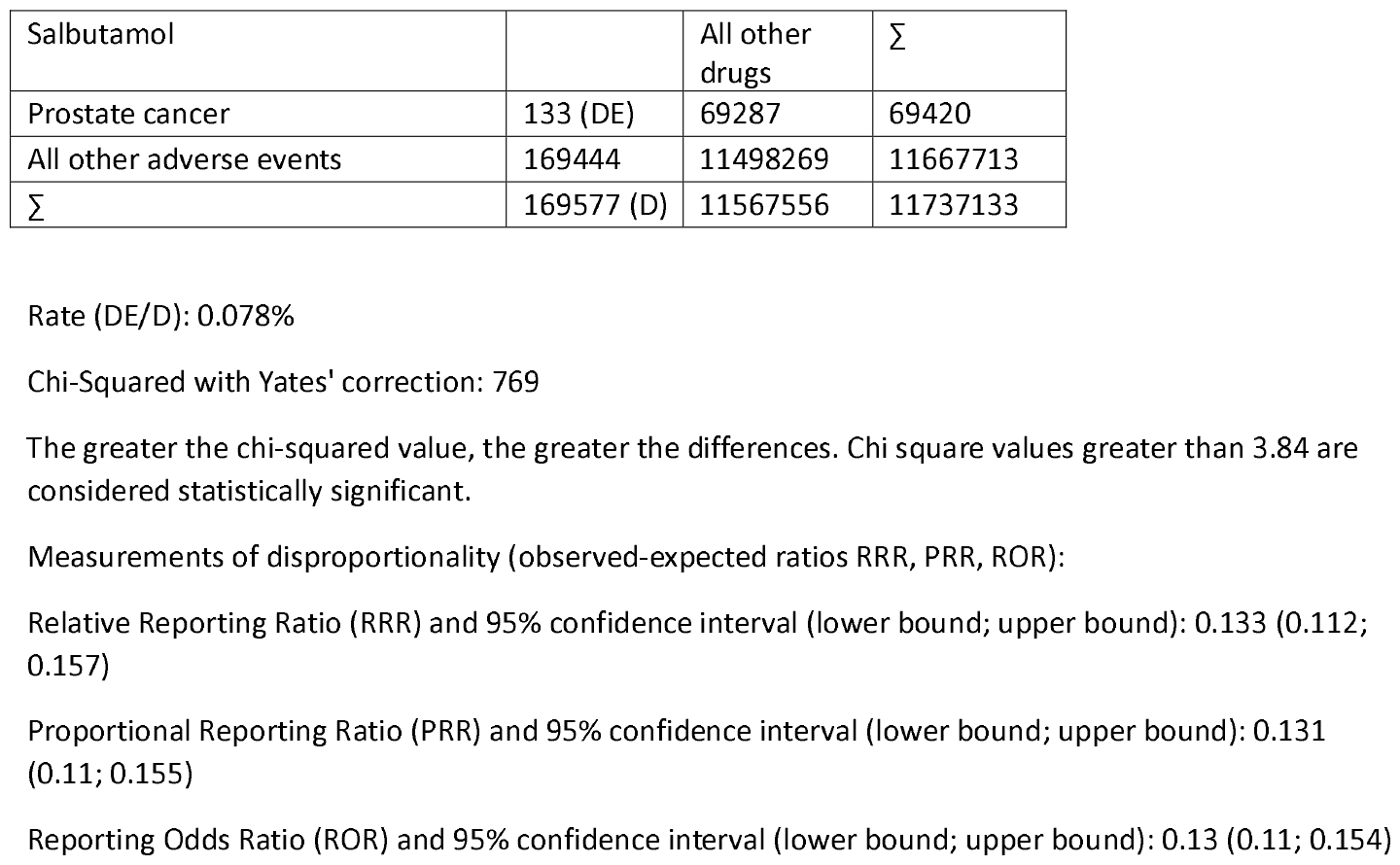
MedWatch data to evaluate the relationship of salbutamol use to prostate cancer in 11,737,133 subjects. Salbutamol use is associated with a significantly reduced risk of prostate cancer PRR (0.131).

Figure 1 flowchart demonstrates how cases and controls were chosen from the full population of unrelated white Europeans in UK Biobank. Figure 2 shows survival probability of prostate cancer patients stratified by salbutamol use. Salbutamol significantly increased survival probability (p = 0.041, log rank test). Mean survival was 7.35 years for subjects not taking salbutamol, 10.5 years for subjects taking salbutamol.

**Figure 1.**
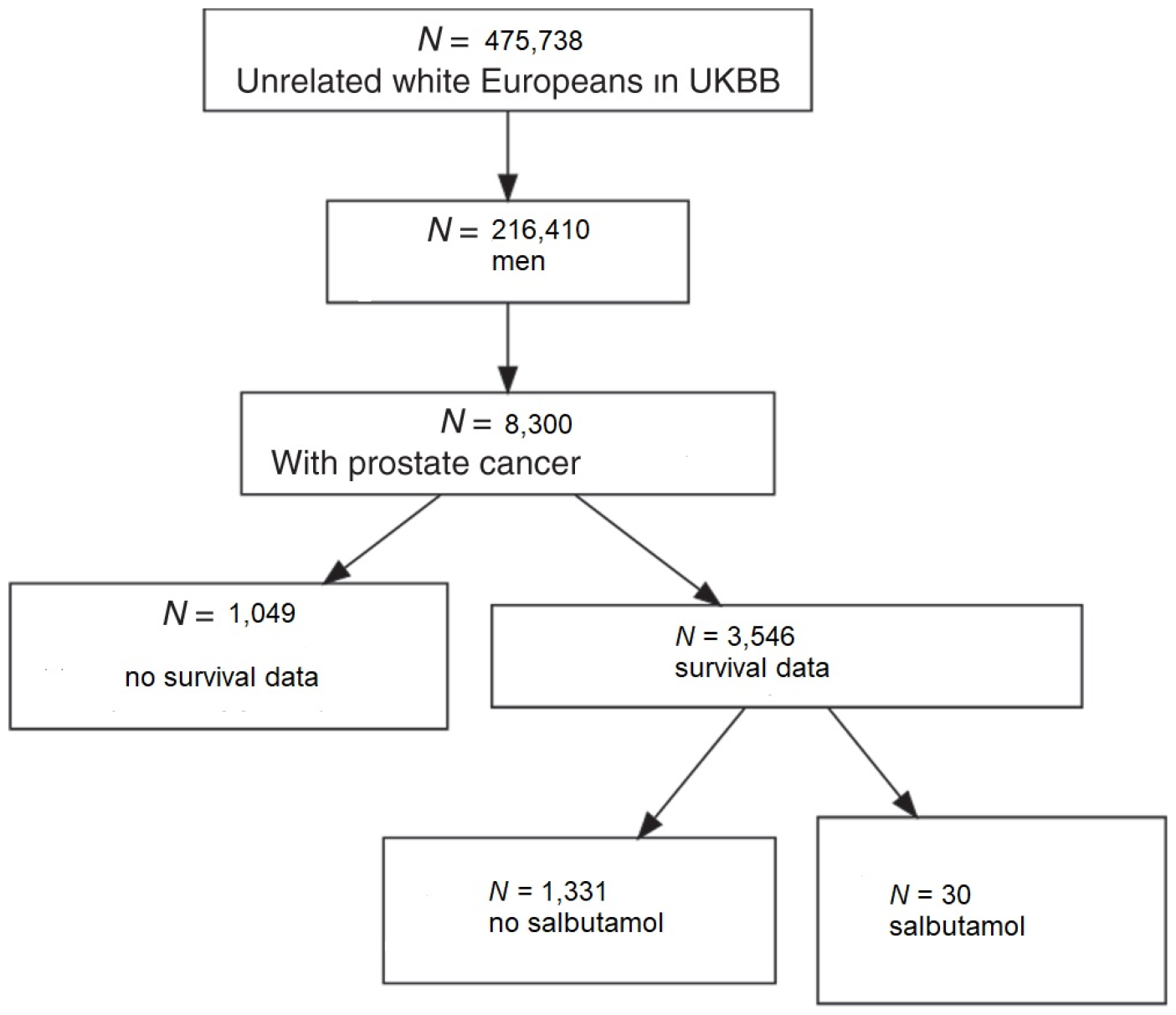
Flowchart demonstrating how cases and controls were chosen from the full population of unrelated white Europeans in UK Biobank. N = number.

**Figure 2.**
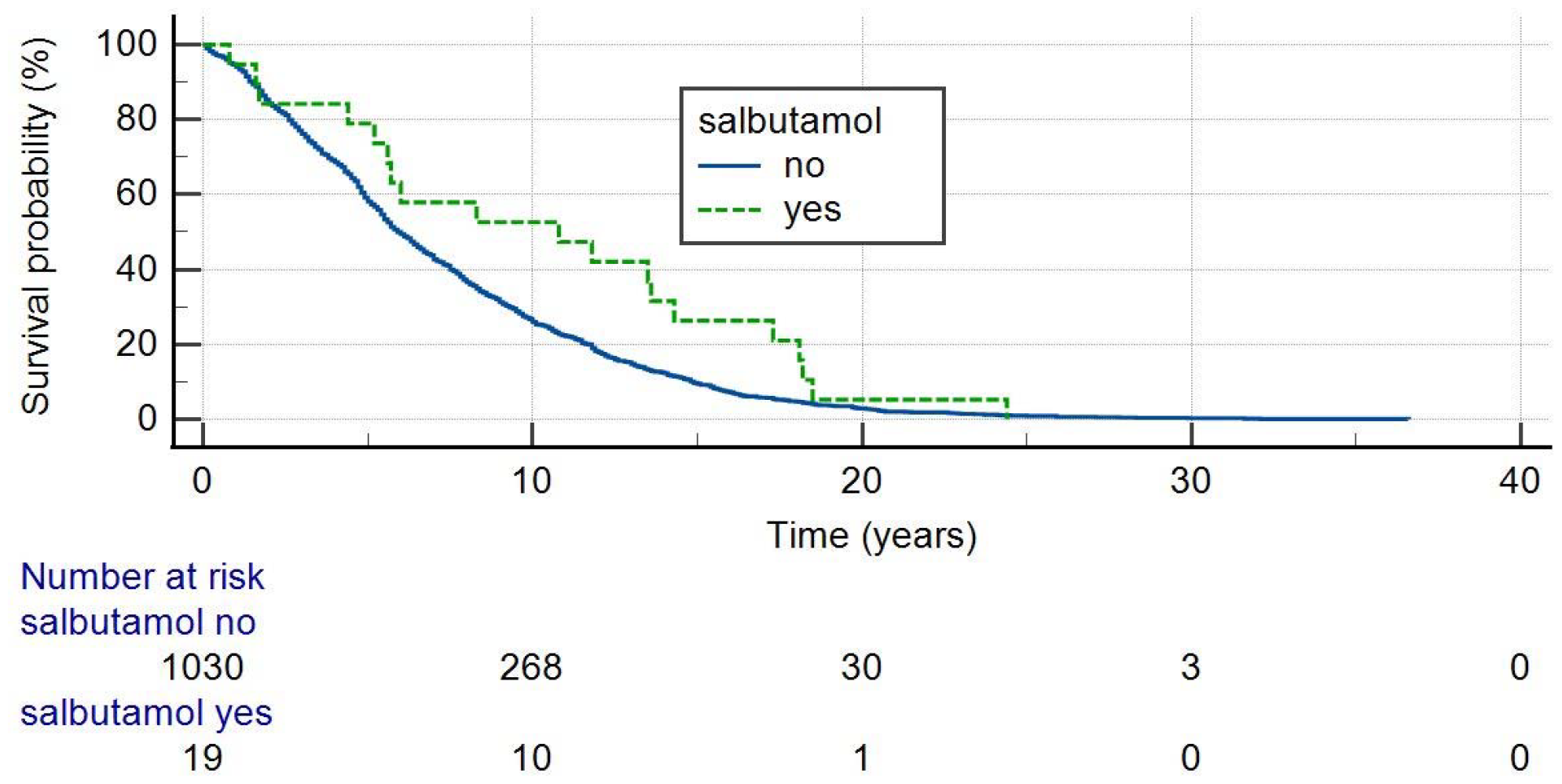
Survival probability of prostate cancer patients stratified by salbutamol use. Salbutamol significantly increased survival probability (p = 0.041, log rank test).

To adjust for the effect of age we performed proportional hazards regression, survival time dependent variable, age and salbutamol use independent variables. Salbutamol use was significantly related to survival time (p = 0.016) and independent of the significant effect of age (p < 0.001).

## Discussion

Beta-2 agonists were developed to treat asthma by modifying the epinephrine molecule. These modifications allow selective interaction with beta-2 receptors on bronchial smooth muscle, leading to bronchodilation. Unlike epinephrine, beta-2 agonists cause less tachycardia because they do not significantly activate beta-1 receptors on cardiac muscle. Activation of beta-2 receptors triggers the enzyme adenylyl cyclase, resulting in the production of cyclic adenosine monophosphate (cAMP). The exact mechanism of smooth muscle relaxation via cAMP is not fully understood but likely involves changes in intracellular calcium concentrations and activation of protein kinase A. Additionally, beta-2 agonists affect potassium channels through a separate mechanism. Beta agonists exhibit varying onset and duration of bronchodilation, depending on their structure. For instance, long-acting beta-agonists (LABAs) like formoterol and salmeterol have long, lipophilic side chains that enhance their binding duration to adrenergic receptors. These modifications contribute to prolonging the bronchodilator effect compared to isoproterenol. Short-acting, selective beta-2 adrenergic agonists such as salbutamol are a mainstay for acute asthma therapy. Long-acting, selective beta-2 adrenergic agonists, often combined with inhaled glucocorticoids, play a role in long-term control of moderate to severe asthma. In addition, beta agonists are utilized in the treatment of chronic obstructive pulmonary disease (COPD) (11,12).

The neuroscience of cancer is involved with the intricate relationship between cancer and the nervous system. In prostate cancer, so-called nervous system hijacking occurs. Just as tumors recruit blood vessels to feed themselves, they also rely on the nervous system for various functions. Prostate cancer cells hijack neural signals to initiate and spread. This co-opting of the nervous system is essential for tumor survival and progression. Sympathetic nerves help during the early stages of prostate cancer development, and parasympathetic nerves boost later-stage spread. Beta blockers disrupt this process (13). Salbutamol might also interfere through neural desensitization and downregulation.

The catecholamines epinephrine and norepinephrine activate beta adrenergic receptors. But these receptors can be desensitized and downregulated by overstimulation. In conditions like heart failure (HF), sustained sympathetic adrenergic activation can lead to beta adrenergic receptor desensitization/downregulation in the myocardium. This process diminishes adrenergic signaling and cardiac contractile reserve, which is conventionally considered detrimental in HF progression (14). Pharmacologic doses of salbutamol over a prolonged period could also, therefore, desensitize and downregulate beta adrenergic receptors in the prostate, reducing prostate cancer risk and improving prognosis.

### Our study has multiple weaknesses

A MedWatch report of an adverse event does not establish causation. For any given report, there is no certainty that the drug in question is related to the reaction. The adverse event may have been due to the underlying disease being treated, another drug being taken concurrently, or something else. The MedWatch data are imperfect, with under- and over-reporting, missing denominator (that is, number of doses for a drug), wrong, duplicate and/or missing data in the database (15). Consequently, the total number of adverse event reports for all drugs and/or the drug in question from OpenVigil can vary slightly from drug to drug and for different adverse events related to the same drug. The imperfect MedWatch data have presented a problem that all analytical software programs, such as OpenVigil, have been forced to confront (16).

UK Biobank does not have PSA level, tumor stage, or Gleason score for prostate cancer, and so we were unable to include these parameters in our analysis.

In people with asthma or COPD, corticosteroids are frequently administered along with salbutamol and other beta-adrenergic agonists. In metastatic, castration resistant prostate cancer, corticosteroids are widely used to palliate symptoms, but are associated with worse outcome (17). Therefore, corticosteroid use could be distorting the results of our study, although diminished PC risk and improved prognosis should be even more significant without corticosteroids.

In conclusion, we have found that salbutamol reduces prostate cancer risk and improves prognosis. Salbutamol and other beta-adrenergic agonists could represent a new class of drugs for treatment of prostate cancer. Further studies are warranted.

## Data Availability

Data sources described in the article are publicly available or can be accessed after approved application to UK Biobank.

https://www.ukbiobank.ac.uk/

http://h2876314.stratoserver.net:8080/OV2/search/

